# ADHD and family life: a cross-sectional study of ADHD prevalence among pupils in China and factors associated with parental stress

**DOI:** 10.1101/2023.01.19.23284771

**Authors:** Tao Lv, Gerard Leavey, Longlong Li, Ying Tang

## Abstract

**Background:** Attention Deficit Hyperactivity Disorder (ADHD) is increasingly recognized as a major problem for children and their families in China. However, its influence on parental stress has been seldom explored.

**Objective:** To examine the prevalence of attention deficit hyperactivity disorder in a community sample of children aged 6-13 years, and the extent to which it impacts parental depression.

**Method:** Cross-sectional study of primary school pupils (number=2497) in Deyang, Sichuan Province, South-West China. We used standardized instruments to identify children with ADHD symptoms and parent depression.

**Results:** The prevalence of ADHD was 9.8%. Factors associated with the likelihood of ADHD, included family environment(P=0.003), time spent with children(P=0.01), parenting style(P=0.01), and parental relationship, pupils self-harm and lower academic ability (P=0.001). After controlling for other factors, having a child with ADHD increased the likelihood of parents’ depression (OR=4.35, CI=2.68∼7.07), additional factors included parent relationship.

**Conclusions:** ADHD may be a common disorder among Chinese children, the symptoms of which may increase the likelihood of parent depression. There is a need for greater detection of ADHD in schools and an acknowledgement of the challenges the disorder creates for academic success and family wellbeing.

## Introduction

Attention deficit hyperactivity disorder (ADHD) is a common developmental and behavioral disorder with the following core symptoms: inattention, hyperactivity, and impulsivity[1]. Learning difficulties, oppositional defiant disorder, anxiety, depression, and other disorders are frequently associated with these symptoms[2]. According to a recent study, the prevalence of ADHD among children aged 6 to 17 was 9.5% in the United States[3], apparently higher than the global prevalence rate of around 5%[4]. There have been no national epidemiological surveys of ADHD in China, and prevalence statistics are not reported uniformly across China’s regions[5]. According to a meta-analysis in 2018, 5.6% of Chinese children are living with ADHD[6]. The overall prevalence of ADHD in China is 6.3% [7]. Furthermore, the economic costs of ADHD are considerable[8]. ADHD symptoms have a significantly negative impact on academic performance and lives[9]. Thus, ADHD patients have high rates of contact with the criminal justice system than the general population[10], causing significant problems for patients, families, and society[11]. Moreover, parents of children with ADHD have a higher prevalence of depression, anxiety disorder, and addictive disorder[12].

In China, less than 2% of children with ADHD seek medical servicer^[13]^ with serious long-term consequences [14,15]. Compared to parents of non-ADHD children, mothers of ADHD children exerience considerable emotional and mental health problems[16,17,18,19,20]. It has been suggested that when parental stress is intensified, such parents may adopt a strict parenting style, which in turn, exacerbates the children’s behavioral issues[20]. Commonly, parents believe that ADHD-related behavior is intentional, resulting in poor parent-child interactions and potential exacerbation of ADHD symptoms[21].However, little is known about the influence of family factors and their relationship with children who exhibit ADHD behaviours. Evidence suggests that compared to their western counterparts, Chinese parenting may be more authoritarian than their Western counterparts[22]. In the West, authoriatrian parenting is regarded negatively as it is associated with challenging behaviors and adjustment problems. However, cultural factors may produce quite different outcomes in Western and Chinese contexts^[23,24]^. Thus, an authoritarian parenting style in China appears to produce acadmic success for children who are expected to be obedient, listen to adults and conform to group expectations, intended to promote respect for others and nurture useful social skills^[25]^.

While evidence suggests that parenting styles may influence behaioural outcomes for chilren with ADHD, there have been no large, community-based studies in mainland China to consider the relationship between ADHD symptoms, family factors, parenting styles and psyhological stress.

## Aims

To assess the likely prevalence of ADHD among schoolchildren in an urban setting in China.To examine the independent impact of children with ADHD symptoms on parental mental health adjusting for other relevant factors.

## Materials and Methods

### Participants

We used Stratified cluster sampling, randomly selecting two primary schools from each urban district, and recruiting 14 schools in Deyang, Sichuan Province, in China. Students in grades 1-6 were randomly screened at each school. Teachers in charge of classes in all schools received standardized training to understand the symptoms of ADHD. Parents and guardians also received assistance in completing the scale. The survey ran from April 2021 to December 2021.

### Demographic questionnaire

Sociodemographic questions included children’s sex, parents’ age, education level, household family income, marital status, household composition, and parental relationships. Parents also reported their children’s academic ability (per school statement). We also collected parents’ time spent with children (hours), parenting style (The parent’s own perception of the family’s upbringing), pupils harm themselves (Parents know this behavior).

### SNAP-IV 26-Item Teacher and Parent Rating Scale[26]

The SNAP scale was developed in accordance with the diagnostic criteria for attention deficit hyperactivity disorder (ADHD) in the DSM (Diagnostic and Statistical Manual of Mental Disorders and has been widely used for ADHD screening, auxiliary diagnosis, and treatment efficacy evaluation in children and adolescents aged 6 to 18 years old. It has three subscales: attention deficit, hyperactivity-impulsion, and oppositional disobedience, with a 0–3-point scale and four grades. Chinese translation reference

### The Patient Health Questionnaire-9, PHQ-9[27]

PHQ-9 is used for rapid screening and symptom assessment of depressive symptoms and consists of nine questions with a total score of 0-4 points for no depression, 5-9 points for mild depression, 10-14 points for moderate depression, 15-19 points for moderate to severe depression, and 20-27 points for severe depression. It has high reliability and validity in detecting major depressive episodes and assessing the severity of depressive symptoms^[28]^.

Ethics

The Ethics Committee of People’s Hospital of Deyang provided ethical approval for research protocol (2021-04-041-K1).

### Data analysis

Data were analyzed using SPSS 28.0 (IBM Corp., Armonk, NY, USA). We used simple descriptive statistics to examine the sociodemographic characteristics of the pupil population, including the proportion with ADHD symptoms and likelihood of ADHD. The chi-square test was performed to compare differences between categorical variables and outcomes of interest. We examined parent’s depression at different levels of severity; first all participants scoring >4 on the PHQ-9; then scores >6 (mild to severe); then scores >10 (severe). We used logistics regression to determine the relationship between ADHD symptoms and specific explanatory variables. We also used logistic regression to examine factors independently associated with first the wider and the second, more restricted parameters of parental depression. The LR results are reported using Odds Ratios and 95% Confidence Intervals.

## Results

### General Demographics

A total of 2513 questionnaires were distributed, of which 46 parents declined to complete, giving a response rate of 98%. This questionnaire was completed by parents or grandparents, (Mothers = 77.6%). Pupils were aged 6-13 years (M=8.51, SD=1.83). Males (n=1060) comprised 42.5% of the sample.

### Prevalence of ADHD

Almost a tenth (9.8%) had ADHD symptoms. Boys were more likely than girls to have ADHD symptoms (11.3% v 8.7%; P=0.05). Child self-harm was also associated with ADHD. ADHD was more common among children with lower academic performance with a noticeable gradient effect (p=0.001). Parent education was associated with symptoms with more fathers in the highest and lowest educated categories (9.4% and12.4%, respectively). A higher proportion of remarried families (23%) had a child with ADHD symptoms than other categories (P=0.003) and poor parental relationships were also significantly associated with child ADHD (24% compared to 7% of those with a good relationship, p=0.001). Those parents with a strict parenting style had a higher proportion of children with ADHD compared to liberal or democratic styles (P=0.01). More hours spent by parents with their children had a modest association with ADHD. (Table 1).

**Table 1.**
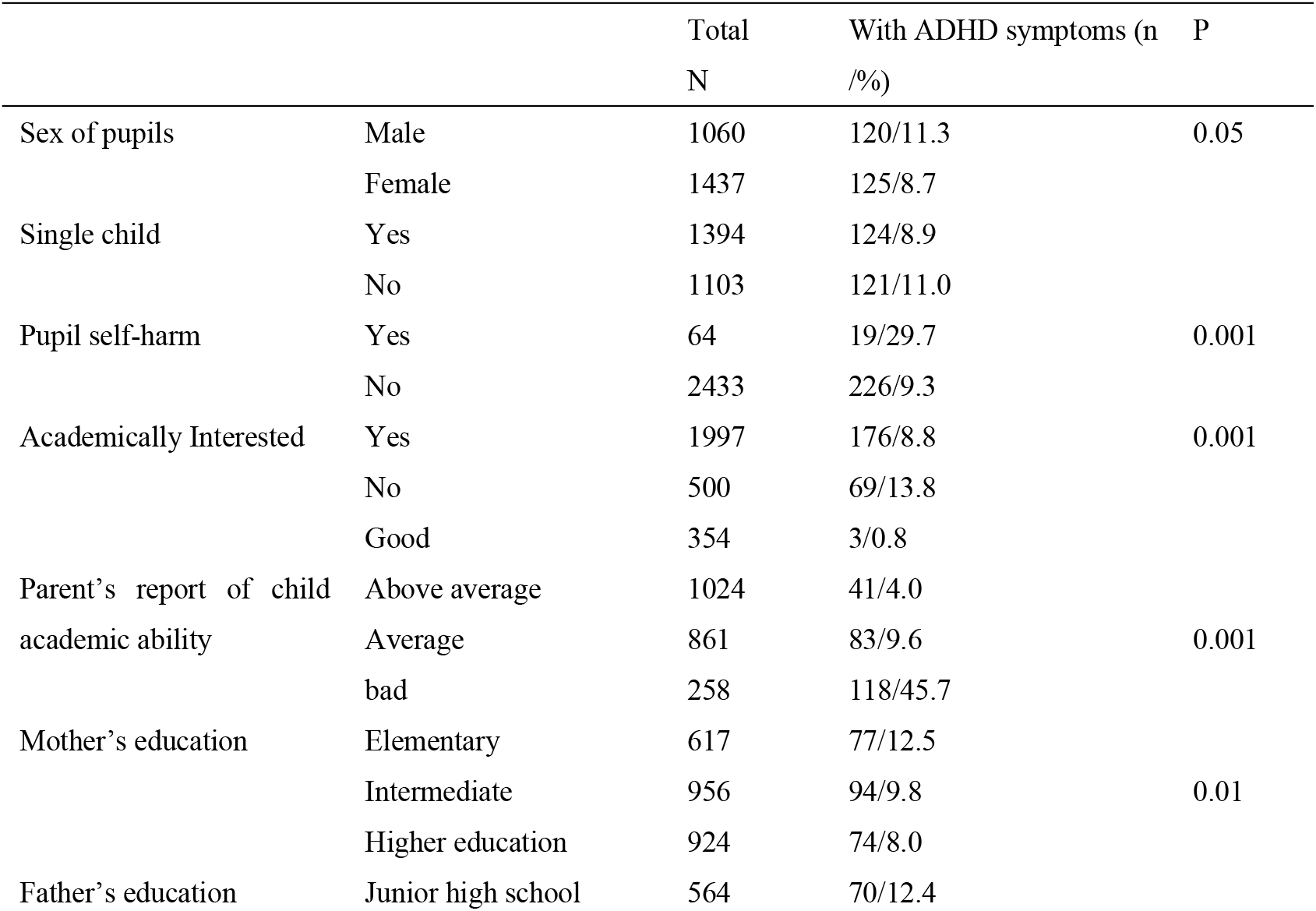
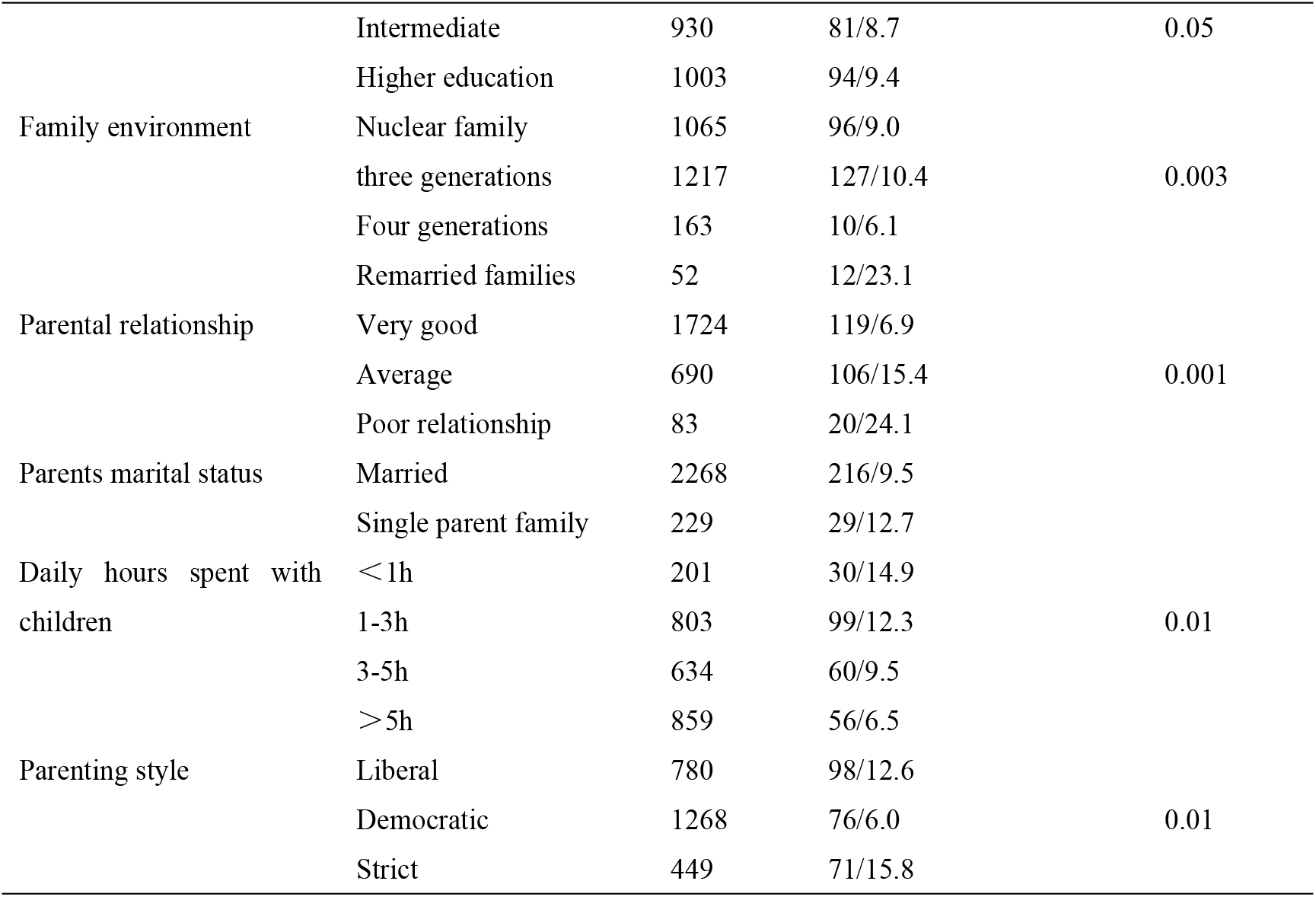
univariate analysis of Pupil ADHD and family context factors(X^2^)

Logistic regression analysis, controlling for annual household income, parents’ education, single parent status, and sex of child indicated that risk of ADHD symptoms were independently associated with child self-harm (OR=3.02, CI=1.66-5.48); remarried families (OR=2.30, CI=1.10-4.84); less hours spent with children. ADHD was associated with a threefold risk among those who reported a “poor marital relationship”, compared to those whose relationship was stated as “good”. Lastly, there was a consistent relationship between parent reported pupil academic performance, with higher assessment and decreasing risk of ADHD. (Table 2).

**Table 2.**
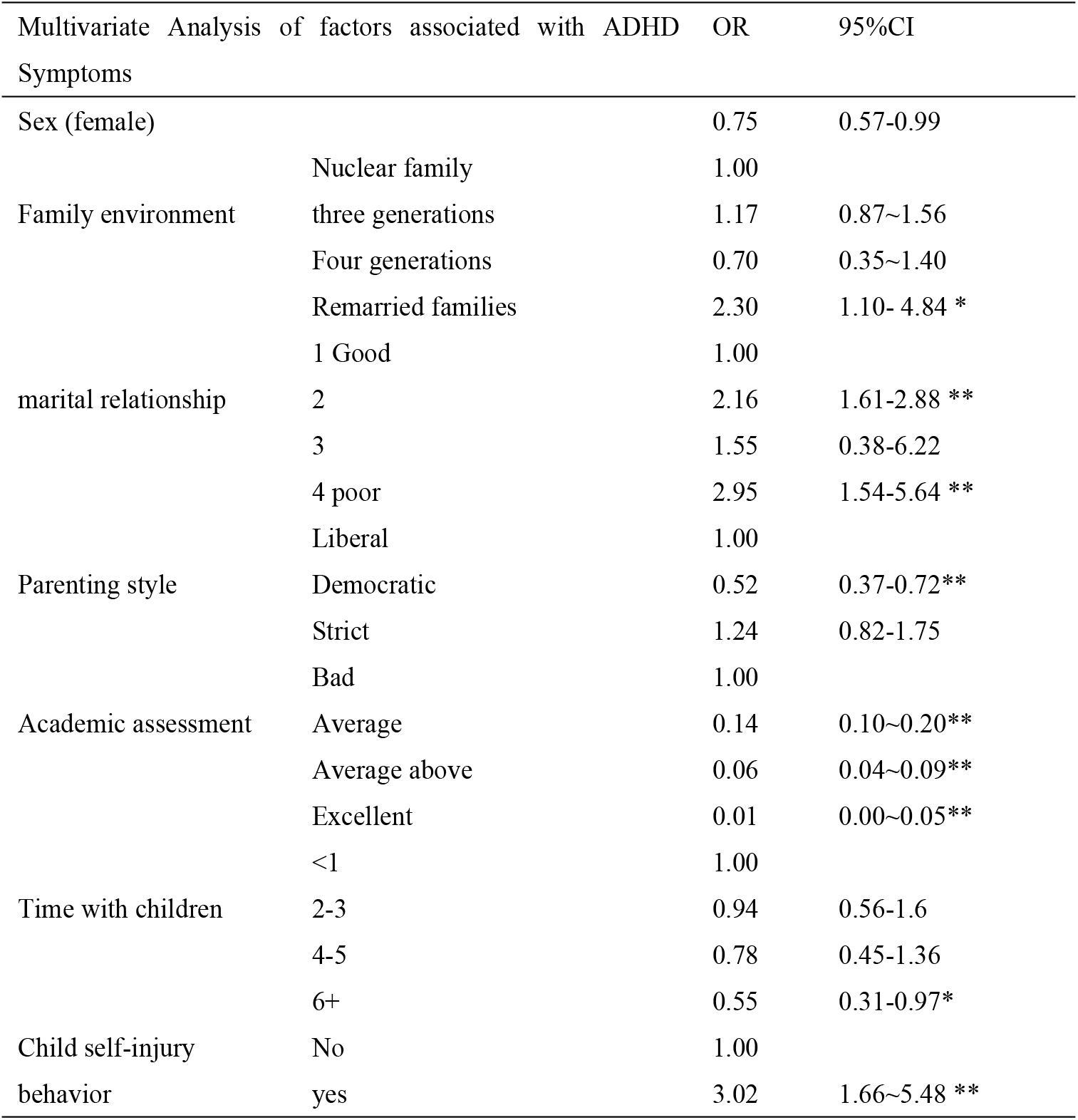
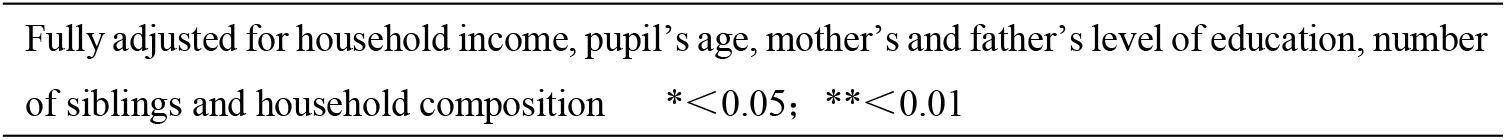
Multivariate analysis of factors associated with ADHD symptoms

| Multivariate Analysis of factors associated with ADHD Symptoms |  | OR | 95%CI |
| --- | --- | --- | --- |
| Sex (female) |  | 0.75 | 0.57-0.99 |
| Family environment | Nuclear family | 1.00 |  |
|  | three generations | 1.17 | 0.87~1.56 |
|  | Four generations | 0.70 | 0.35~1.40 |
|  | Remarried families | 2.30 | 1.10- 4.84 * |
| marital relationship | 1 Good | 1.00 |  |
|  | 2 | 2.16 | 1.61-2.88 ** |
|  | 3 | 1.55 | 0.38-6.22 |
|  | 4 poor | 2.95 | 1.54-5.64 ** |
| Parenting style | Liberal | 1.00 |  |
|  | Democratic | 0.52 | 0.37-0.72** |
|  | Strict | 1.24 | 0.82-1.75 |
| Academic assessment | Bad | 1.00 |  |
|  | Average | 0.14 | 0.10~0.20** |
|  | Average above | 0.06 | 0.04~0.09** |
|  | Excellent | 0.01 | 0.00~0.05** |
| Time with children | <1 | 1.00 |  |
|  | 2-3 | 0.94 | 0.56-1.6 |
|  | 4-5 | 0.78 | 0.45-1.36 |
|  | 6+ | 0.55 | 0.31-0.97* |
| Child self-injury behavior | No | 1.00 |  |
|  | yes | 3.02 | 1.66~5.48 ** |

Fully adjusted for household income, pupil's age, mother's and father's level of education, number of siblings and household composition \* $<0.05$ ; \*\* $<0.01$

### Parent Depression

Using a cut-off of 4 or more on the PHQ, approximately 28% of the parents had some form of depression (mild-severe). More restricted classifications of (>6) indicated that 22% (n=546) had moderate to severe depression, while scores >10 indicated 4.8% of people (n=120) had severe depression. In bivariate analysis, 55% of people who reported a “poor marital relationship” had some level of depression which was less common among those who reported joint caregiving (among mothers and fathers). Depression was more common among those who reported that their children had self-harmed (58% v 27%, P=0.001) poor academic achievements for their child (P=0.001); strict or liberal parenting styles (P=0.001). Annual household income had a modest and unclear relationship with depression. (Table 3).

**Table 3:**
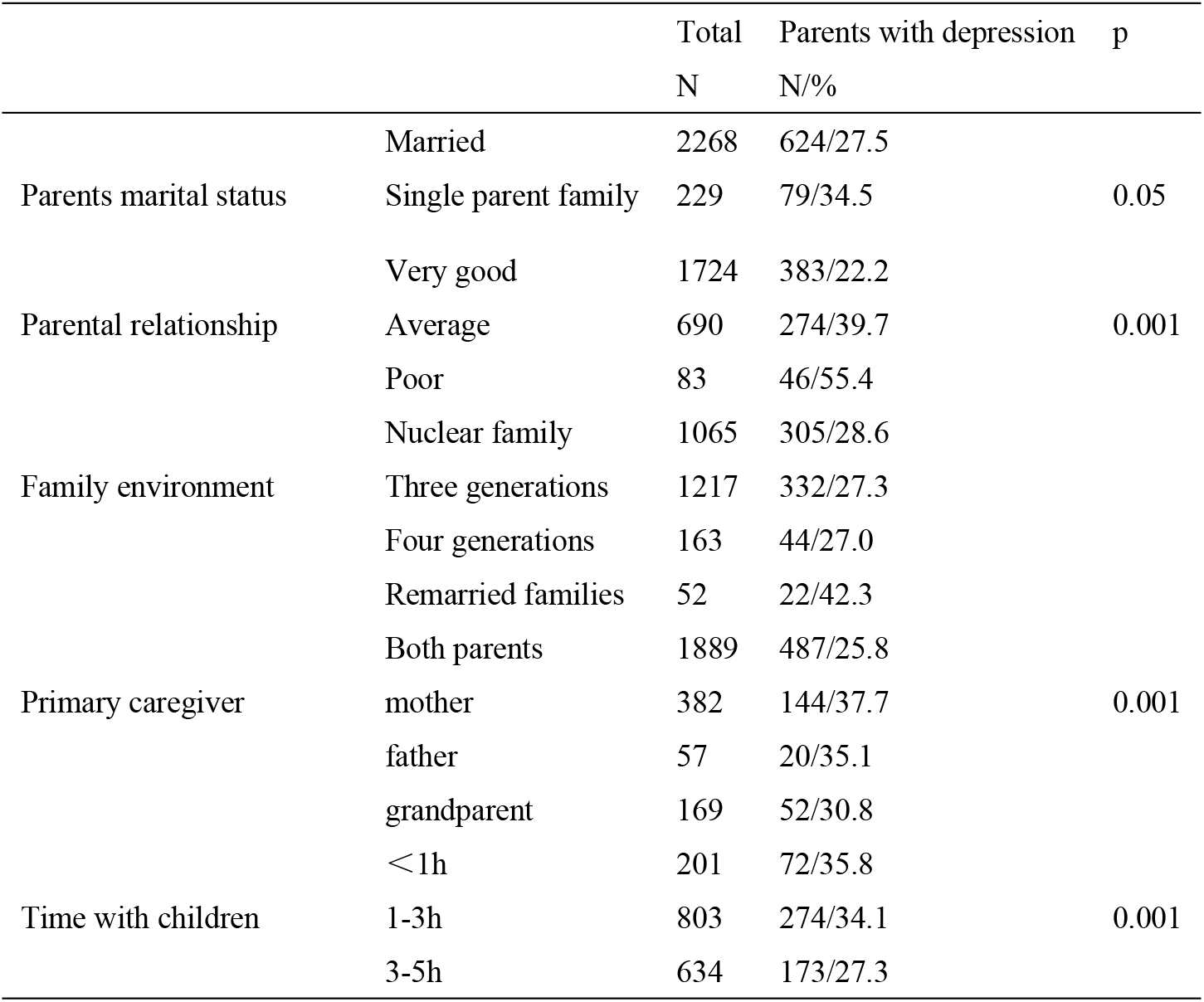
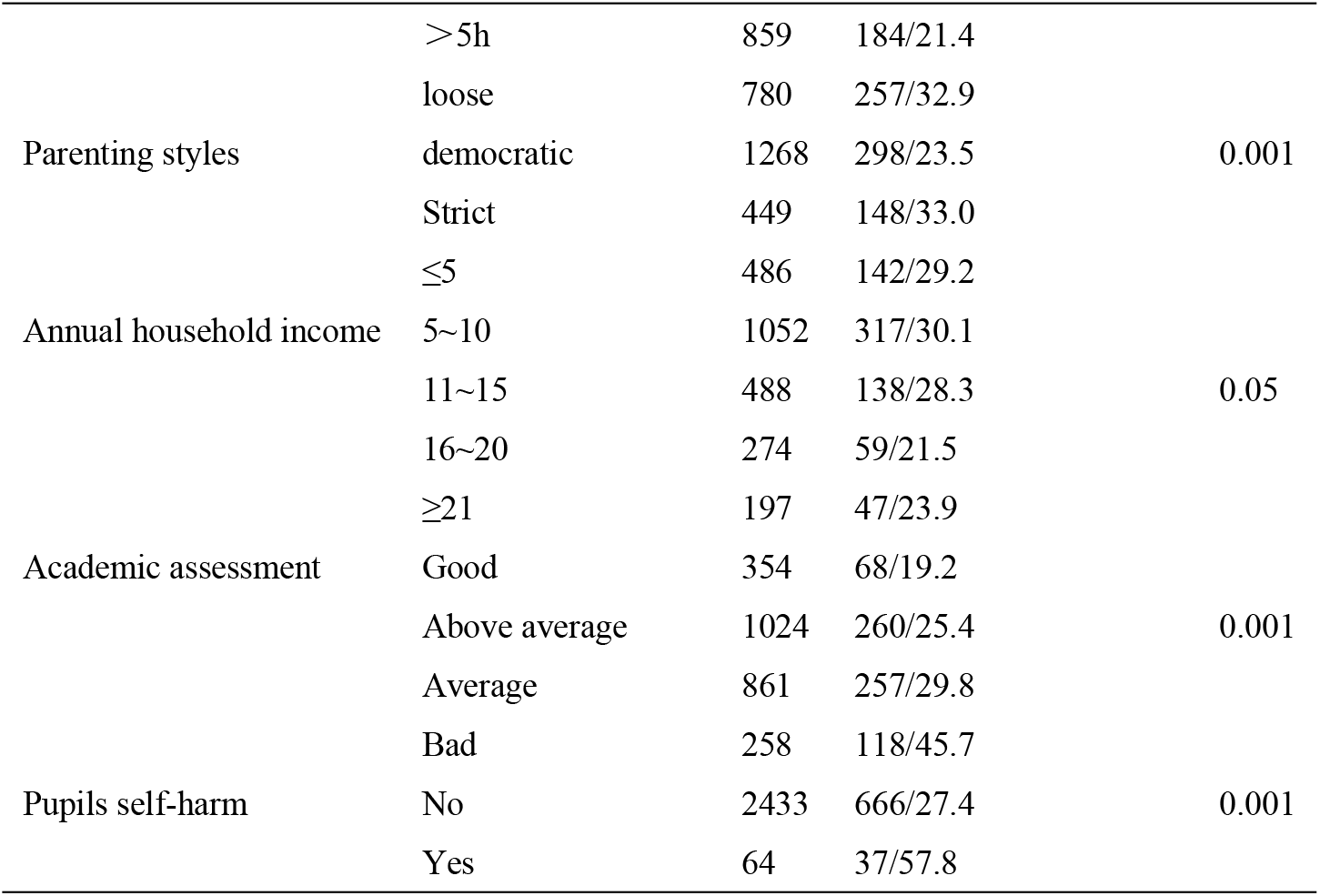
Univariate family life factors associated with parental depression (X^2^)

### Factors associated with parental depression

Using a binary logistic regression, we examined factors related to parental depression. First, we examined all cases (anyone scoring >6) on the PHQ. Families with one child had an increased risk of depression but of borderline significance. Compared to families where child caregiving is evenly shared, mothers (not fathers) who had the primary caregiving duties were at increased risk of depression (OR=1.62, CI=1.22-2.15). The amount of time spent with children is somewhat protective; significantly so for parents who spent more than 5 hours per day (OR=0.64, CI=0.41-0.98). Parents with a ‘democratic” parenting style, had a reduced likelihood of pression, compared to parents who reported a strict parenting style (OR=0.70, CI=0.56-0.89). A liberal parenting style also had a reduced but non-significant risk. Participants who reported relationship difficulties were significantly more at risk of depression, compared to those who reported harmonious relationships (OR=3.46, CI=2.00-6.00). Those with children who self-harmed had a greater likelihood of depression but fell short of significance. Lastly, parents whose child scored positive for ADHD were more at risk of depression (OR=2.73, CI=2.00-3.72). Our multivariate analysis of parent depression using a higher threshold of >10 on the PHQ, indicated that mothers recorded as having the main caregiving responsibilities had an increased likelihood of depression (OR=2.40, CI=1.48-3.90). Those who spent more than 5 hours per day with their children had a significantly reduced risk of depression. Reported child self-harm and ADHD symptoms both increased the likelihood of parental depression. Parenting styles, parents’ educational levels, and reported pupil’s academic ability lost statistical significance. (Table 4)

**Table 4.** Multivariate analysis of factors associated with parental depression at higher and lower levels of severity (for PHQ scores >6; and scores >10)

|  | Parents with depression scores<br>greater than 6 (n=546, 22%) |  |  | Parental depression Scores<br>greater than 10 (n=120, 4.8%) |  |  |
| --- | --- | --- | --- | --- | --- | --- |
|  | OR | 95%CI |  | OR | 95%CI |  |
|  |  | Lower | Upper |  | lower | Upper |
| **Single child** | 1.03 | 0.83 | 1.27 | 0.94 | 0.63 | 1.42 |
| **Primary Care** |  |  |  |  |  |  |
| Both parents | 1.00 |  |  |  |  |  |
| Only mother | **1.62** | **1.22** | **2.15 \*\*** | 2.40 | 1.48 | 3.90 |
| Only father | 1.25 | 0.61 | 2.54 | 0.86 | 0.21 | 3.48 |
| Grandparent | 1.27 | 0.82 | 1.20 | 0.83 | 0.35 | 2.01 |
| **time with children (hours)** |  |  |  |  |  |  |
| <1 | 1.00 |  |  |  |  |  |
| 2-3 | 0.91 | 0.60 | 1.37 | 0.90 | 0.44 | 1.81 |
| 4-5 | 0.77 | 0.50 | 1.19 | 0.62 | 0.30 | 1.32 |
| >5 | **0.64** | **0.41** | **0.98 \*** | **0.27** | **0.11** | **0.62 \*\*** |
| **Parenting styles** |  |  |  |  |  |  |
| Strict | 1.00 |  |  | 1.00 |  |  |
| Democratic | **0.70** | **0.56** | **0.89 \*\*** | 0.88 | 0.55 | 1.41 |
| Liberal | 0.90 | 0.68 | 1.20 | 1.35 | 0.81 | 2.24 |
| **Family environment** |  |  |  |  |  |  |
| Nuclear | 1.00 |  |  |  |  |  |
| 3 generations | 0.89 | 0.72 | 1.10 |  |  |  |
| 4 Generations | 1.03 | 0.67 | 1.58 |  |  |  |
| Remarried | 1.28 | 0.67 | 2.44 |  |  |  |
| **Pupil self-Harm (yes)** | 1.73 | 0.99 | 2.10 | **2.27** | **1.08** | **4.77\*** |
| **ADHD symptoms (yes)** | **2.73** | **2.00** | **3.72 \*\*** | **4.35** | **2.68** | **7.07 \*\*** |
| **parent relationship** |  |  |  |  |  |  |
| 1 Good | 1.00 |  |  |  |  |  |
| 2 | **1.88** | **1.51** | **2.23 \*\*** | **1.50** | **0.99** | **2.30** |
| 3 | **4.00** | **1.41** | **11.36 \*\*** | **2.90** | **0.69** | **12.71** |
| 4 Poor | **3.46** | **2.00** | **6.00 \*\*** | **3.74** | **1.72** | **8.10 \*\*** |
| **marital status** | 0.84 | 0.56 | 1.27 |  |  |  |
| **father education** | **1.21** | **1.03** | **1.43 \*** |  |  |  |
| **mother education** | 1.12 | 0.95 | 1.33 |  |  |  |
| **Academic assessment** | 0.85 | 0.74 | 0.96 * | 0.95 | 0.74 | 1.22 |

Controlling for age and sex of children, annual household income P= > 0.05\* p=>0.001 \*\*

## Discussion

The global prevalence of ADHD is around 5%[4], and according to the most recent research, the prevalence of ADHD among children aged 6-17 in the United States has reached 9.5%[3]. The prevalence surveys are inconsistent across regions of China. We found a prevalence of ADHD of almost 10% among these pupils living in seven urban area of De yang district of China, higher than those recorded in other Chinese studies[29]. Boys had a higher prevalence than girls, consistent with other findings[6]. Unlike Huang et al[30], we noted that families with more than one child had a higher incidence rate than those with single children.

We also observed that reported child self-harm was more common among children with ADHD. Importantly, our findings highlight the severe impact of ADHD and academic performance as reported by the parents. This is consistent with previous evidence about the social and educational challenges related to behavioral symptoms of this population (e.g., poor concentration, fidgeting,). It may also be the case that parents who find their children’s’ behaviors as problematic, may also negatively rate their child’s academic abilities. However, parental assessment was based on that provided by their school report and this may be a more reliable and objective statement. Children and young people with ADHD have a higher risk of academic and social problems, and conflicts with pupils and teachers[31].

We also found that ADHD was related to remarried families, again suggesting that the challenges of parenting children with ADHD may present a higher risk of marital breakdown, noted in previous studies[32]. Perhaps related, ADHD was associated with a three-fold risk in those who reported a poor parental relationship. It is possible that parents whose relationships are problematic reproduce behavioral and psychological problems in their children, and this may explain the higher levels of recorded ADHD symptoms in such children. However, most of the evidence suggests that family breakdown is more likely to result in depression and anxiety, rather than the ADHD symptoms recorded in this study. Moreover, we found that parental relationships posed an independent risk for parental depression, after controlling for the independent effects of child self-harm. ADHD was also associated with an authoritarian parenting styles, and fewer hours spent with their children. Previous evidence suggests that maternal psychopathology (particularly depression) can predict poor treatment outcomes in ADHD patients[33].

### Parental depression

In our regression analysis of factors related to parental depression, we examined the likelihood of ADHD symptoms exerting an independent effect on parent psychological distress. We did this in a robust examination of restricted thresholds for depression and found that ADHD had an increased and independent effect on parental depression. Thus, parents with children who scored positive for ADHD had more than a four-fold risk of depression at at the lower and higher scores on the PHQ (10+ OR=4.35, CI=2.68-7.07). Unsurprisingly, child self-harm was also independently associated with a higher risk of parental depression. Importantly, we noted a protective effect where both parents are noted as being joint caregivers; mothers with sole caregiving responsibilities were more likely to be depressed (OR=1.62, CI=1.22-1.67). It is important to note that in China, (as in this sample), many households had three or four generations living together but this does not appear to reduce maternal stress and may contribute to it through additional responsibilities, and household income appears to have little influence on wellbeing. Increasingly in China, many families are now dual-career families, unable to balance work and life, with less time to accompany children[34].

Parental depression appears strongly related to ADHD symptoms in their children, consistent with previous research^[35]^. Lastly, compared to parents who endorsed a more authoritarian (strict) parenting style, those who endorsed “liberal” or “democratic” styles were less likely to be depressed but this was significant only for “democratic” and scoring greater than six on the PHQ (OR=0.70, CI=0.56-0.89). It may be that while parenting style has an independent effect on parental depression, many parents adopt this style as an approach to coping with ADHD symptoms in their children but that this may personally uncharacteristic, and consequently heightens stress and depression.

Active screening of parents’ emotions and children’s ADHD symptoms can serve as the foundation for future family guidance. Improving children’s ADHD symptoms can reduce the likelihood of parental depression and improving parents’ depression can also promote the alleviation of ADHD symptoms. Family support should be strengthened in clinical practice to promote the improvement of parent-child relationships and the quality of family life.

## Limitations

We found a higher prevalence of ADHD symptoms than in other studies using clinical interview[30], but similar results to other international surveys[36]. However, it should also be acknowledged that cultural differences may play a part in the assessment of ADHD symptoms and behaviors^[30]^. Secondly, our study would have benefited from corroborative data in addition to that provided by parents. However, it is more likely that parents would under-report challenging behaviors due to stigma[37]. Third, while most of our instruments were well-validated for a Chinese context, we were obliged to use self-designed or adapted questions such as those relating to parenting styles.

## Conclusions

ADHD may be a common disorder among Chinese children, the symptoms of which may increase the likelihood of parent depression. There is a need for greater detection of ADHD in schools and an acknowledgement of the challenges the disorder creates for academic success and family wellbeing.

## Data Availability

The data underlying the results presented in the study are available on request from Professor Leavey at

## Acknowledgments

we want to thank all the participating schools and teachers who helped makes this study possible. Additional thanks to all the parents and grandparents who gave their time.

